## Supplementary information for "Individual level analysis of digital proximity tracing for COVID-19 in Belgium highlights major bottlenecks"

---

Authors: Caspar Geenen, Joren Raymenants, Sarah Gorissen, Jonathan Thibaut, Jodie McVernon, Natalie Lorent, Emmanuel André

### Supplementary Tables

---

***Supplementary table 1. Test indications and their description in the online test booking form.*** *Test indications evolved over the duration of the test and trace program, in line with changing national guidelines. All test indications used throughout the extended study period are listed here with their identifier codes, implementation dates and exact wording in the English version of the online test booking form.*

| **Identifier** | **Period implemented** | **Description in test booking form** |
|---|---|---|
| pos_self_test | from 27/04/2021 | a positive self test |
| symptoms | before 25/08/2021 | immediately after getting mild COVID-19 symptoms |
| | from 25/08/2021 | mild COVID-19 symptoms (as soon as possible) |
| close_contact | before 25/08/2021 | immediately (between 1 and 3 days) after a high-risk contact with a proven COVID-19 positive person |
| | | 7 days after a high-risk contact with a proven COVID-19 positive person |
| | 25/08/2021 - 16/12/2021 | a risk contact (preferably on days 1 to 3, with a second test on day 7) |
| | from 16/12/2021 | a risk contact (see website for advised test dates) |
| return_abroad | before 27/04/2021 | on day 1 after returning from a red zone abroad |
| | | on day 7 after returning from a red zone abroad |
| | | on day 14 after a return from or passing through India, Latin America or South Africa |
| | from 25/08/2021 | arrival from a red zone (on days 1 and/or 7, depending on government's instructions after PLF (passenger locator form) |
| coronalert | before 25/08/2021 | after getting a red screen on the CoronAlert app |
| | from 25/08/2021 | a red screen on CoronAlert |
| extended_tracing | before 25/08/2021 | by referral from the contact tracers of the KU Leuven who informed me about a confirmed infection in my student residence |
| | from 25/08/2021 | referral from the contact tracers of the KU Leuven who informed me about a confirmed infection |
| start_internship | throughout study period | the start of my (para)medical internship |
| precaution | before 25/08/2021 | no specific reason |
| | from 25/08/2021 | a concern about a possible infection |
| medical_treatment | from 25/08/2021 | a hospital admission or treatment, that requires preventive testing |

***Supplementary table 2. Model input parameters for comparing manual or digital contact tracing to case isolation only.*** *This table lists the parameters used to model the effect of different TTIQ strategies (test, trace, isolate, quarantine) on the effective reproduction number in our setting. The parameters were used as input for the tti package in R[1,2]. Baseline: no intervention. Isolation: case isolation only. DPT: case isolation with DPT. MCT: case isolation with MCT. DPT+MCT: case isolation with combined DPT and MCT*

| **Parameter** | **Explanation** | **Input value** | **Source** |
|---|---|---|---|
| alpha | The probability of an asymptomatic infection | **0.2** | [3] |
| R | Effective reproduction number in fully vaccinated Delta/Omicron dominant population with moderate contact restrictions | **1.5** | The national $R_{eff}$ (based on case numbers) started at 1.15 at the start of the main study period (18 October 2021), climbed to 1.28 at the start of November, decreased to a minimum of 0.69 at the start of December. With the arrival of omicron variant, it increased again to a new peak of 1.68 at the end of December, before declining again to 1.15 at the end of the study period[4]. The $R_0$ associated with the Delta and Omicron variants imply that a baseline $R_{eff}$ of 1.5 or 2 can only be achieved with a combination of immunity, general contact restrictions, and barrier measures[5,6]. |
| Kappa | Relative transmissibility of an asymptomatic individual compared to a symptomatic individual. | **0.35** | [3] |
| Eta | Probability that a contact is a household contact. | **0** | We do not differentiate between household and non-household contacts in this model. |
| nu | Relative risk of infection for a household contact compared to a community contact. | **1** | Not relevant, as we do not consider household contacts separately. |
| t_ds | Time delay from symptom onset to isolation in detected symptomatic person. [days] | Baseline: **100**<br>Other strategies: **2.34** | Sum of the mean delays between (1) symptom onset and sampling, and (2) sampling and PCR test result. (Supplementary Figure 4). |
| t_da | Time delay from symptom onset to isolation in detected asymptomatic person. [days] | **100** | Practically infinite delay, as we considered only symptomatic screening as index case detection strategy. |
| t_qcs / t_qhs | Time delay from symptomatic index cases's symptom onset to quarantine of | Baseline: **100**<br>Isolation: **100**<br>MCT: **4.60**<br>DPT: **3.52** | Sum of t_ds and estimate of the mean delay from case PCR test result to contact notification. This delay is |

| Parameter | Explanation | Input value | Source |
|---|---|---|---|
| | community/household contacts. [days] | MCT+DPT: **4.49** | determined for MCT by fitting a log-normal curve to the distribution of observed values. For DPT, it is determined with a log-normal curve as described in *Methods*. The delay for a combined DPT and MCT strategy is determined by calculating the probability that either a manual or digital notification is received, based on both lognormal curves. |
| t_qca / t_qha | Time delay from asymptomatic index cases's symptom onset to quarantine of community/household contacts. [days] | **100** | We considered only symptomatic screening as index case detection strategy. |
| t_q | Time delay from quarantined index cases's symptom onset to quarantine of contacts. [days] | Equal to t_qcs | Assumed equal to detection through symptomatic screening. |
| omega_c / omega_h / omega_q | The probability of being traced and quarantined given community / household / quarantine contact of a person. | Baseline: **0**<br>Isolation: **0**<br>MCT: **0.441**<br>DPT: **0.043**<br>MCT+DPT: **0.464** | As per the estimation in *Table 1*. No differentiation by contact type. |
| quarantine_ days | The number of days contacts are told to quarantine. [days] | **7** | Standard guidance in Belgium at the time. |
| rho_s | The probability of detection and isolation given symptomatic. Product of case acertainment and effectiveness of isolation to interrupt transmission (assumed to be 100). | Varied from **0.1 to 0.9** in 0.1 increments | Highly reliant on specific testing resources and willingness to test[7–9]. |
| rho_a | Same as rho_s, but for asymptomatic individuals. | **0** | We considered only symptomatic screening and contact tracing as case detection strategies. |
| t_incubation | Mean incubation period. [days] | **4.2** | Average of incubation periods of the Delta and Omicron BA1 variants[10]. |
| offset | Offset of infectiousness compared to symptoms onset. [days] | **-2.31** | Baseline model assumption[1], based on [11]. |
| shape | Shape of the gamma distribution of infectious period. | **1.65** | Baseline model assumption[1], based on [11]. |
| rate | Rate of the gamma distribution of infectious period. | **0.5** | Baseline model assumption[1], based on [11]. |

| Parameter | Explanation | Input value | Source |
| --- | --- | --- | --- |
| stoch | Whether to run stochastic model with overdispersion. | **FALSE** | |
| Theta | Overdispersion parameter for negative binomial distribution. | **0.1** (only for sensitivity analysis with stochastic model) | Baseline model assumption[1], based on [11]. |
| N_inf | Number of infections (or effective population size of infected individuals). | **20** (only for sensitivity analysis with stochastic model) | Common number of new daily infections in the university testing centre |
| N_iter | Number of iterations of stochastic model to run for each unique parameter value. | **100,000** (only for sensitivity analysis with stochastic model) | |

***Supplementary table 3. Results of sensitivity analyses when modelling the effective reproduction number.*** *Selected parameters were altered relative to the nominal model parameters in Supplementary table 2. This table lists which parameters were altered and the resulting modelled effect of each contact tracing strategy on the effective reproduction number, relative to case isolation only (columns 2 to 4). The last two columns show the effect of DPT and combined DPT/MCT, respectively, on the effective reproduction number, relative to MCT (with case isolation also implemented in each strategy).*

| **Altered parameter(s)** | **Case isolation + DPT** | **Case isolation + MCT** | **Case isolation + DPT + MCT** | **Effect of DPT relative to MCT (given case isolation)** | **Effect of DPT+MCT relative to MCT (given case isolation)** |
|---|---|---|---|---|---|
| Model without alterations | 1.06 | 1.58 | 1.63 | 0.11 | 1.08 |
| Stochastic model | 1.06 | 1.62 | 1.68 | 0.09 | 1.09 |
| Stochastic model,<br>N_iter = 200,000 | 1.06 | 1.62 | 1.67 | 0.10 | 1.08 |
| Lower limit (2.8%) of DPT success confidence interval, i.e.,<br>multiply all omega values with 0.65. | 1.04 | 1.37 | 1.39 | 0.11 | 1.08 |
| Upper limit (6.1%) of DPT success confidence interval, i.e.,<br>multiply all omega values with 1.42. | 1.09 | 1.87 | 1.95 | 0.10 | 1.09 |
| Lower limit (0.6) of DPT delay confidence interval, i.e.,<br>t_qcs, t_qhs, t_q =<br>2.97 (DPT)<br>4.45 (DPT+MCT) | 1.07 | 1.58 | 1.64 | 0.11 | 1.09 |
| Upper limit (2.2) of DPT delay confidence interval, i.e.,<br>t_qcs, t_qhs, t_q =<br>4.57 (DPT)<br>4.56 (DPT+MCT) | 1.05 | 1.58 | 1.62 | 0.09 | 1.07 |
| 4-day delay from case PCR result to MCT notification, i.e.,<br>t_qcs = t_qhs = t_q =<br>6.34 (MCT)<br>6.07 (DPT/MCT) | 1.06 | 1.35 | 1.41 | 0.17 | 1.18 |
| alpha = 0.5 | 1.06 | 1.61 | 1.66 | 0.10 | 1.08 |
| alpha = 0.1 | 1.06 | 1.58 | 1.63 | 0.11 | 1.08 |
| R = 1.2 | 1.06 | 1.58 | 1.63 | 0.11 | 1.08 |
| R = 5 | 1.06 | 1.58 | 1.63 | 0.11 | 1.08 |
| Kappa = 2 | 1.06 | 1.62 | 1.67 | 0.10 | 1.08 |
| Kappa = 0.2 | 1.06 | 1.58 | 1.63 | 0.11 | 1.08 |
| t_ds = 4 (all except baseline strategy) | 1.15 | 2.46 | 2.57 | 0.10 | 1.08 |

| | | | | | |
|---|---|---|---|---|---|
| t_ds = 1 (all except baseline strategy) | 1.03 | 1.25 | 1.27 | 0.11 | 1.09 |
| Add 1 day to values of t_qcs, t_qhs and t_q | 1.05 | 1.46 | 1.50 | 0.11 | 1.10 |
| Subtract 1 day from values of t_qcs, t_qhs and t_q | 1.07 | 1.68 | 1.73 | 0.10 | 1.07 |
| Multiply all omega values with 0.5 | 1.03 | 1.28 | 1.30 | 0.11 | 1.08 |
| Multiply all omega values with 1.5 | 1.09 | 1.93 | 2.01 | 0.10 | 1.09 |
| t_incubation = 2 | 1.06 | 1.53 | 1.58 | 0.12 | 1.10 |
| t_incubation = 6 | 1.05 | 1.50 | 1.54 | 0.10 | 1.08 |
| offset = 0 | 1.01 | 1.08 | 1.08 | 0.10 | 1.09 |
| offset = -4 | 1.15 | 2.36 | 2.48 | 0.11 | 1.09 |

### Supplementary Figures

***Supplementary Figure 1. Exclusion chart for the infection risk analysis by test indication for the main study period (18 October 2021 - 9 January 2022).*** *See Methods for the exact questions asked, and Supplementary table 1 for a full list of test indications implemented throughout the study period and their grouping for this analysis.*

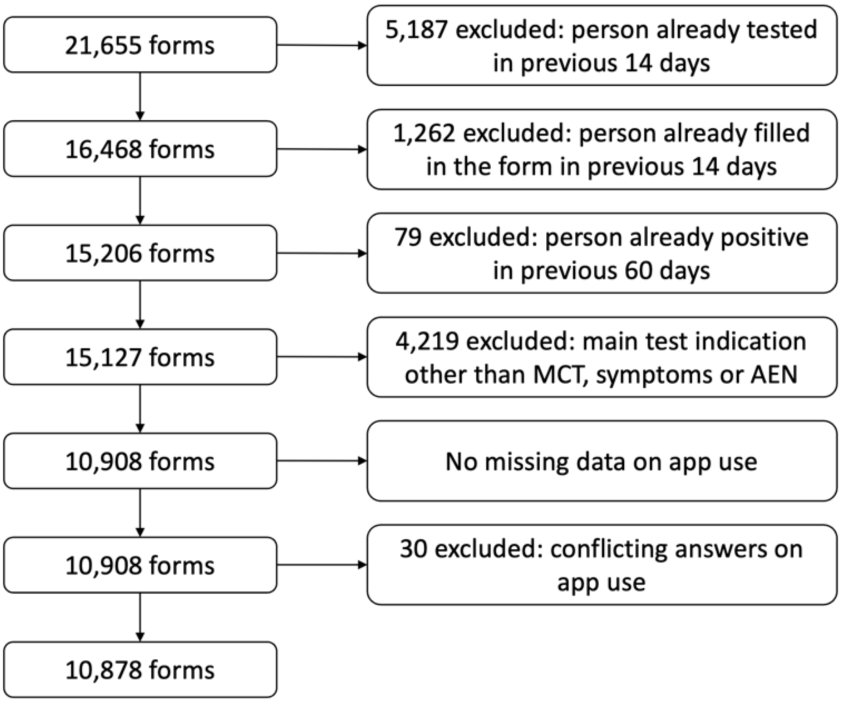

***Supplementary Figure 2. Exclusion chart for the infection risk analysis by test indication for the extended study period (1 February 2021 until 21 March 2022).*** *See Methods for the exact questions asked, and Supplementary table 1 for a full list of test indications implemented throughout the study period and their grouping for this analysis.*

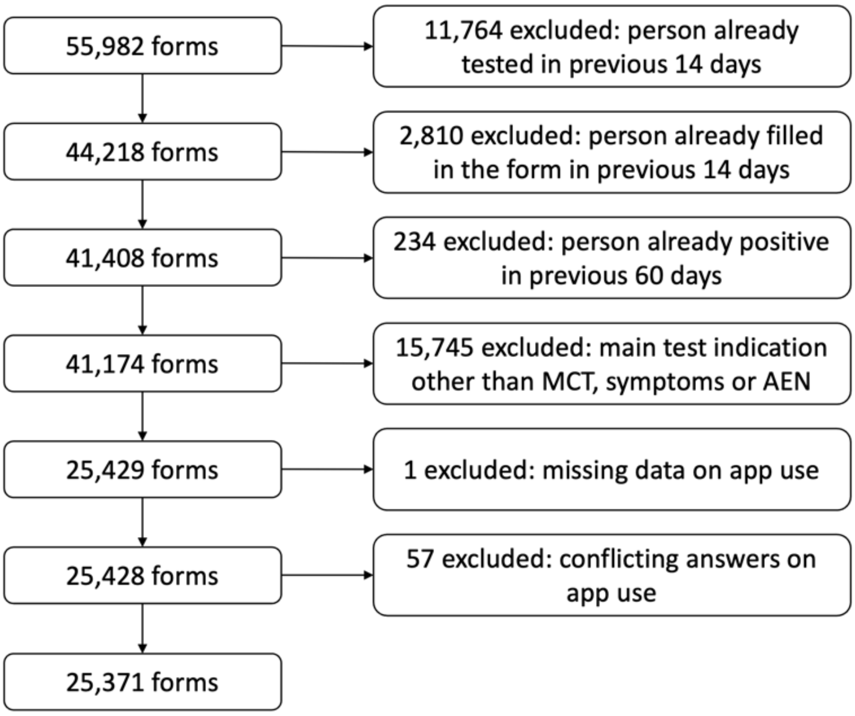

***Supplementary figure 3. Number of tests, infection risk and risk ratio by self-reported test indication for persons undergoing a test at the university test centre in the extended study period (1 February 2021 - 21 March 2022).*** *Persons are grouped according to self-reported main test indication: manually traced close contact, receipt of an automated exposure notification (AEN), or symptoms suggestive for COVID-19. The exclusion chart is shown in Supplementary Figure 2. In panel a, each group is subdivided according to app use and AEN receipt. For each subgroup, the number of infected and uninfected persons, the number of persons lost to follow-up, and the infection risk are listed. In addition, the risk ratio is shown relative to non-app users who were manually traced or had suggestive symptoms. In panel b, each group is subdivided according to outcome: infected, not infected or lost to follow-up. For each subgroup, the number of app users with and without an AEN is indicated, as well as the numbers of people not using the app. The rate of AEN receipt amongst app users is shown for each subgroup. Relative risks are calculated compared to a control group of persons attending for suggestive symptoms, but not infected with COVID-19. We consider this control group a surrogate for the average rate of AEN receipt (justified or not) in the population. Reassuringly, both infected symptomatic persons and those with a manually traced close contact had a similarly increased rate of AEN receipt.*

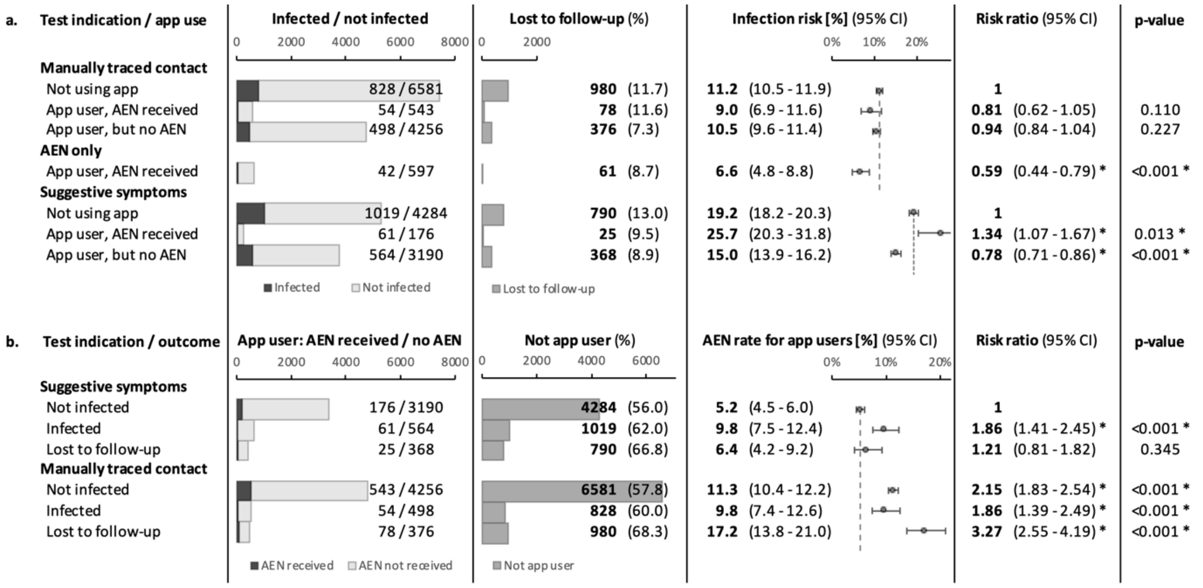

***Supplementary Figure 4. Schematic representation of delays in the digital notification cascade during the main study period.*** *The mean delay is shown for each step, with 95% confidence intervals between brackets. Symptom onset was assessed during contact tracing for persons who were diagnosed through symptomatic screening. The delay between sampling and test result was assessed for persons testing positive at our test centre. Values in italic were modelled by fitting a cumulative log-normal function to the data, as described in Methods and Results.*

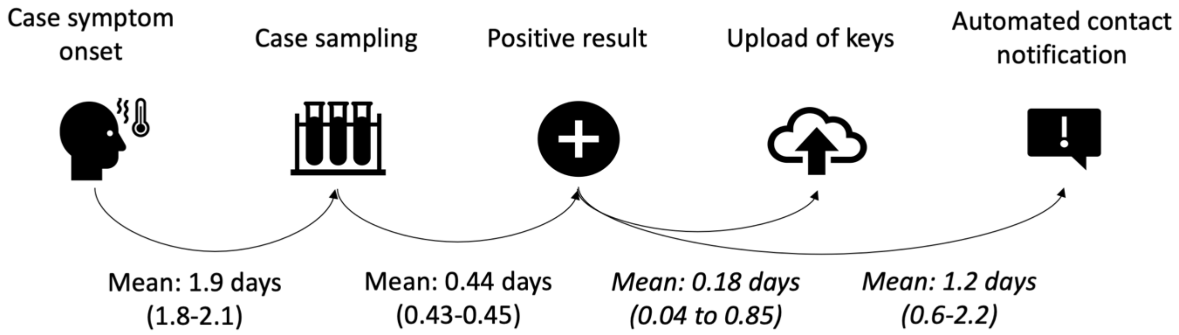

***Supplementary Figure 5. Modelled effect on the effective reproduction number ($R_{eff}$) of different case isolation and contact quarantine strategies in our setting.*** *Panel a shows how the modelled effective reproduction number decreases almost linearly with the proportion of cases identified through symptomatic screening. Panel b shows the effect of each strategy on $R_{eff}$ relative to case isolation only.*

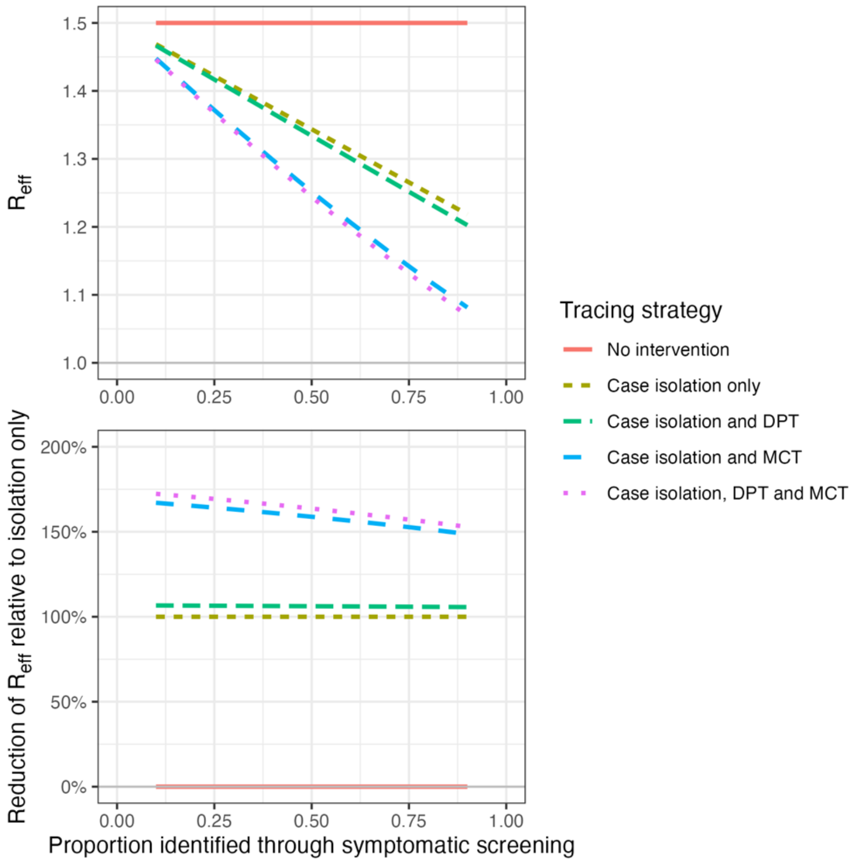

***Supplementary Figure 6. Incidence and additional measures of MCT and DPT performance in the extended study period.*** *Panel a shows the weekly number of confirmed cases amongst Leuven students and the Belgian population. Panels b and d indicate the total number of manually traced contacts and the number of contacts per case, respectively. Panel c shows the proportion of persons who received an automated digital notification (AEN), amongst those attending the test centre for a manually traced exposure. Dots show weekly proportions, and error bars the 95% confidence interval, except in panel d, where grey dots are individual data points. Vertical black lines show the beginning and end of the main study period. Coloured lines show local polynomial regression curves, and the shaded areas their 95% confidence intervals. A change in app configuration, intended to reduce the number of notified contacts, is indicated with a blue vertical line.*

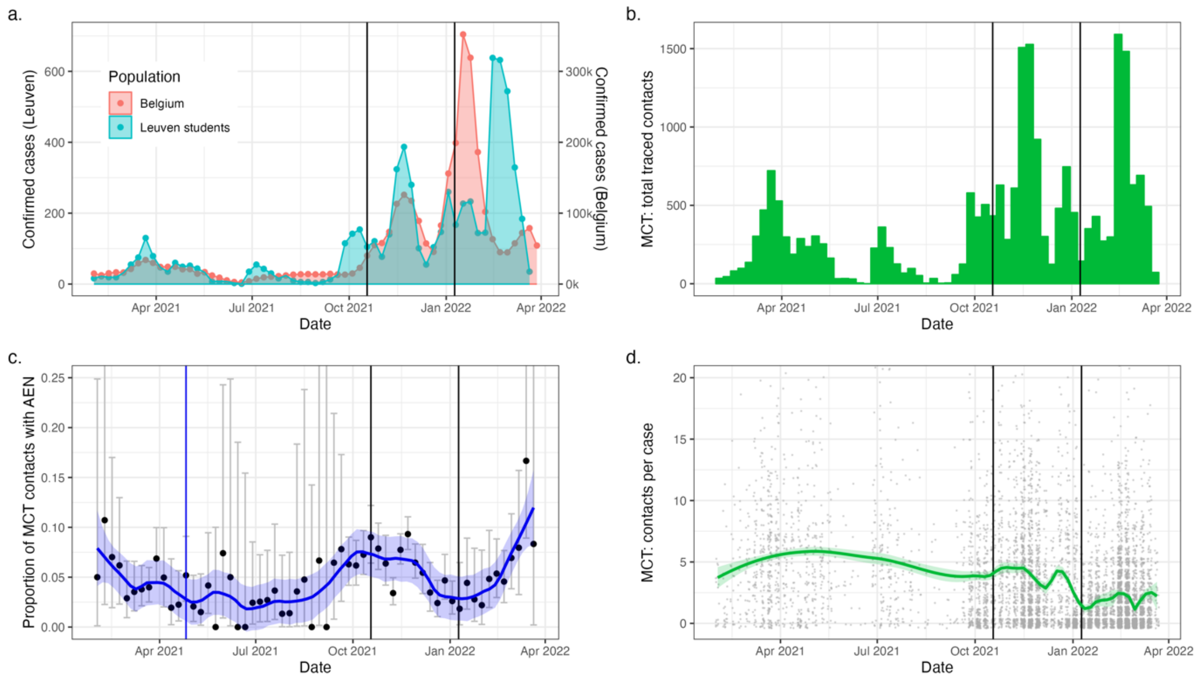
